# Prognostic impact of atrial cardiomyopathy: Long-term follow-up of patients with and without low-voltage areas following atrial fibrillation ablation

**DOI:** 10.1101/2023.06.26.23291924

**Authors:** Masaharu Masuda, Yasuhiro Matsuda, Hiroyuki Uematsu, Ayako Sugino, Hirotaka Ooka, Satoshi Kudo, Subaru Fujii, Mitsutoshi Asai, Shin Okamoto, Takayuki Ishihara, Kiyonori Nanto, Takuya Tsujimura, Yosuke Hata, Naoko Higashino, Sho Nakao, Toshiaki Mano

**Affiliations:** Kansai Rosai Hospital Cardiovascular Center, 3-1-69 Inabaso, Amagasaki, Hyogo 660-0060, Japan

**Keywords:** Low-voltage area, Sex difference, Predictors, Recurrence

## Abstract

**Background:** Atrial cardiomyopathy is known as an underlying pathophysiological factor in the majority of AF patients. Left atrial low-voltage areas (LVAs) are reported to coincide with fibrosis, and to likely represent atrial cardiomyopathy. This study aimed to delineate differences in the long-term prognosis of patients stratified by the size of LVAs.

**Methods:** This observational study included 1,488 consecutive patients undergoing initial ablation for AF. LVAs were defined as regions with a bipolar peak-to-peak voltage of < 0.50 mV. The total study population was divided into 3 groups stratified by LVA size: patients with no LVAs (n=1136), those with small (< 20 cm^2^, n=250) LVAs, and those with extensive (≥ 20 cm^2^, n=102) LVAs. Composite endpoints of death, heart failure, and stroke were followed for up to 5 years.

**Results:** Composite endpoints developed in 105 (7.1%) of 1488 patients, and AF recurrence occurred in 410 (27.6%). Composite endpoints developed more frequently in the order of patients with extensive LVAs (19.1%), small LVAs (10.8%), and no LVAs (5.1%; p for trend<0.0001). Multivariable analysis revealed that LVA presence was independently associated with higher incidence of composite endpoints, irrespective of AF recurrence (modified hazard ratio=1.73, 95% confidence interval=1.13-2.64, p=0.011)

**Conclusions:** LVA presence and its extent were both associated with poor long-term composite endpoints of death, heart failure, and stroke, irrespective of AF recurrence or other confounders. Underlying atrial cardiomyopathy appears to define a poor prognosis after AF ablation.

**Clinical perspective:** *What is Known:* Patients with left atrial low-voltage area has high incidence of atrial fibrillation recurrence after ablation

*What the study adds:* Presence of low-voltage areas was associated with poor prognosis including death, heart failure hospitalization, and stroke.

**Graphic abstract:** 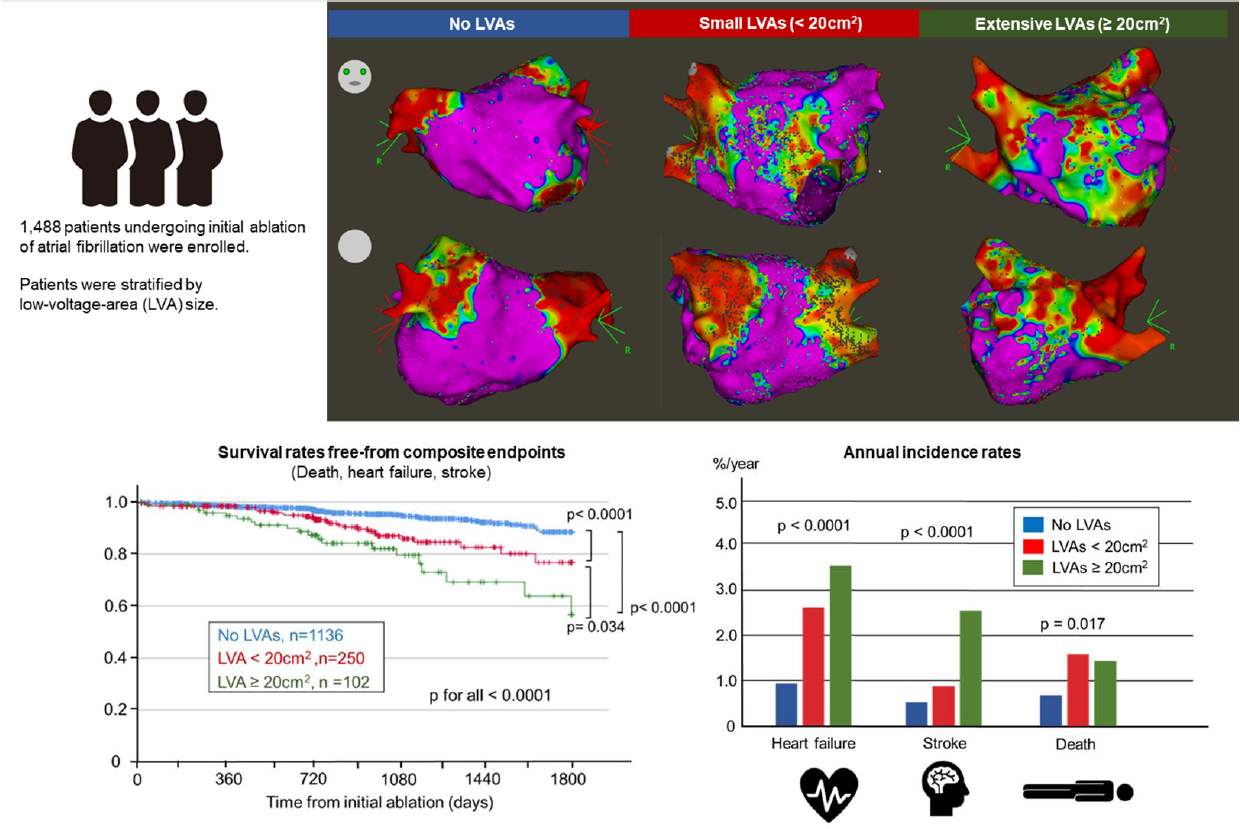

## Introduction

Patients with atrial fibrillation (AF) are known to have a poor prognosis, with increased risk of mortality, heart failure, and stroke.^1^ The term ‘atrial cardiomyopathy’ was initially proposed to describe remodeling of the atria due to upstream factors, genetic disorder, aging, AF burden and others.^2^ Atrial cardiomyopathy is known as the underlying pathophysiology in the majority of AF patients, and to be possibly related to the development of complications of AF such as heart failure and stroke. ^2,3,4^ However, few studies have examined the long-term prognostic impact of atrial cardiomyopathy in AF patients.

Voltage mapping during the AF ablation procedure depicts the distribution of atrial myocardial degeneration, including fibrosis, as low-voltage areas (LVAs).^5^ In the clinical setting, the presence and extent of LVAs have both been shown to correlate well with AF recurrence after AF ablation. ^6,7,8^ In addition, several studies reported that the presence of LVAs is associated with increased atrial stiffness, elevated left atrial pressure, or both.^9,10^ These data suggest that the presence and extent of LVAs are related to atrial remodeling.

Given the lack of a standardized method to assess the severity of atrial cardiomyopathy as background, this study used the size of LVAs as a marker of severity of atrial cardiomyopathy. Our purpose was to delineate differences in long-term prognosis among patients stratified by the size of left atrial LVAs.

## Methods

### Patients

This observational study enrolled 1,488 consecutive patients who underwent initial ablation of AF at Kansai Rosai Hospital from December 2014 to March 2022. Patients with an incomplete left atrial voltage map were excluded. Other exclusion criteria were age < 20 years, prior left atrial catheter or ablation, and prior MAZE surgery. The total study population of 1,488 patients was divided into 3 groups stratified by size of LVAs: patients with no LVAs, those with small (< 20 cm^2^) LVAs, and those with extensive (≥ 20 cm^2^) LVAs. (Central Figure) This study complied with the Declaration of Helsinki. Written informed consent for the ablation and participation in this study was obtained from all patients, and the protocol was approved by our institutional review board.

### Catheter ablation and voltage mapping

Pulmonary vein isolation was performed in all patients using a linear radiofrequency catheter or cryoballoon. Other ablations, including ablation targeting LVAs, linear ablation, ablation for induced atrial tachycardias, and ablation for non-pulmonary vein AF triggers were added at the discretion of the attending physicians. An electroanatomical mapping system (CARTO 3, [Biosense Webster, Inc., Diamond Bar CA, USA]; Ensite NavX, [Abbott, Abbott Park IL, USA]; or Rhythmia, [Boston Scientific, Boston MA, USA]) was used for ablation guidance and mapping.

Following pulmonary vein isolation, left atrial voltage mapping was performed under sinus rhythm or atrial paced rhythm from the right atrium. In most cases, the multi-electrode mapping catheter was a PENTARAY (Biosense Webstar), Lasso NaV (Biosense Webstar), HD grid (Abbott), or Orion (Boston Scientific). A 3.5- or 4.0-mm ablation catheter was used as an auxiliary in some areas in which catheter-tissue contact was difficult. The band pass filter was set at 30 to 500 Hz. Adequate endocardial contact was confirmed by distance to the geometry surface and stable electrograms. Mapping points were acquired to fill all color gaps on the voltage map. Respective fill and color interpolation thresholds were 15 mm and 23 mm for CARTO 3 and 20 mm and 7 mm for Ensite NavX. The interpolation threshold for Rhythmia was set at 5 mm. LVAs were considered present when the summed size of all areas with a bipolar peak-to-peak voltage < 0.50 mV was ≥ 5 cm^2^.^11^

### Follow up for AF recurrence

Patients were followed every 4–8 weeks at the dedicated arrhythmia clinic of our institution for as long as possible, with a minimum of 2 years. Routine ECGs were obtained at each outpatient visit, and 24-h ambulatory Holter monitoring was performed at 6 and 12 months post-ablation. When patients experienced symptoms suggestive of an arrhythmia, a surface ECG, ambulatory ECG, and/or cardiac event recording were also obtained. Either of the following events following the initial 3 months after ablation (blanking period) was considered to indicate AF recurrence: (1) atrial tachyarrhythmia (AF and regular atrial tachycardia) recorded on a routine or symptom-triggered ECG during an outpatient visit; or (2) atrial tachyarrhythmia of at least 30-s duration on ambulatory ECG monitoring. No antiarrhythmic drugs were prescribed after the ablation procedure unless AF recurrence was observed.

### Composite endpoints

Composite endpoints included death from any cause, heart failure, and stroke. Stroke defined as intracranial bleeding and ischemic events were diagnosed by neurologists based on diagnostic imaging such as computed tomography and/or magnetic resonance imaging. Heart failure was defined as symptomatic heart failure requiring hospitalization. Diagnosis of heart failure symptoms was made by cardiologists using the Framingham heart failure diagnostic criteria.^12^ In patients who ceased attending follow up within 5 years after ablation, information on composite endpoints was collected generally by telephone interview.

### Statistical analysis

Continuous data are expressed as the mean ± standard deviation or median (interquartile range). Categorical data are presented as absolute values and percentages. Tests for significance were conducted using the unpaired *t*-test, or nonparametric test (Mann-Whiney *U*-test) for continuous variables, and the chi-squared test or Fisher’s exact test for categorical variables. Cox proportional hazard models were used to determine the prognostic impact of LVAs and other factors. Variables with a p value ≤ 0.05 in the univariate models were included in the multivariate analysis. Event-free survival rates were calculated using the Kaplan-Meier method. Survival curves between groups were compared with the 2-sided Mantel-Haenszel (log-rank) test. All analyses were performed using commercial software (SPSS version 26.0^®^, SPSS, Inc., Chicago IL, USA).

## Results

### Patient characteristics

Patient characteristics in the three groups are compared in Table 1. Patients with LVAs were older, had a smaller body size and weight, were more likely to be female and less likely to have paroxysmal AF, and had a higher brain natriuretic peptide level and lower estimated glomerular filtration rate. Concomitant diseases such as diabetes mellitus, heart failure, and stroke were more frequent in patients with LVAs. Patients with LVAs also had higher clinical scores predicting the presence of LVAs (SPEED and DR-FLASH).^13,14^ On echocardiography, patients with LVAs had a larger left atrial size and similar left ventricular ejection fraction.

**Table 1.** Baseline characteristics.

|  | No LVA<br>*n* = 1136 | LVAs < 20 cm<sup>2</sup><br>*n* = 250 | LVA ≥ 20cm<sup>2</sup><br>*n* = 102 | *p* |
| --- | --- | --- | --- | --- |
| **Age, years** | 66.5 ± 10.4 | 73.3 ± 7.4 | 73.6 ± 6.1 | < 0.0001 |
| **Female, n (%)** | 307 (27.0) | 133 (53.2) | 61 (59.8) | < 0.0001 |
| **Height, cm** | 165 ± 9 | 159 ± 9 | 158 ± 10 | < 0.0001 |
| **Weight, kg** | 66.5 ± 13.3 | 60.0 ± 13.1 | 56.8 ± 12.4 | < 0.0001 |
| **Body mass index, kg/m<sup>2</sup>** | 24.3 ± 3.9 | 23.7 ± 4.6 | 22.6 ± 3.6 | < 0.0001 |
| **AF type** |  |  |  | < 0.0001 |
| *Paroxysmal, n (%)* | 492 (43.3) | 67 (26.8) | 24 (23.5) |  |
| *Persistent AF lasting < 1 year, n (%)* | 552 (48.6) | 163 (65.2) | 67 (65.7) |  |
| *Persistent AF lasting ≥ 1 year, n (%)* | 92 (8.1) | 20 (8.0) | 11 (10.8) |  |
| **Hypertension, n (%)** | 641 (56.4) | 134 (53.6) | 59 (57.8) | 0.668 |
| **Diabetes mellitus, n (%)** | 183 (16.1) | 63 (25.2) | 23 (22.5) | 0.002 |
| **Heart failure, n (%)** | 217 (19.1) | 72 (22.8) | 33 (32.4) |  |
| **Stroke or systemic thromboembolism, n (%)** | 79 (7.0) | 19 (7.0) | 19 (18.6) | < 0.0001 |
| **Vascular disease, n (%)** | 96 (8.5) | 24 (9.6) | 14 (13.7) | 0.19 |
| **CHA<sub>2</sub>DS<sub>2</sub>-VASc score** | 2.3 ± 1.4 | 3.2 ± 1.4 | 3.6 ± 1.4 | < 0.0001 |
| **eGFR, mL/min/1.73m<sup>2</sup>** | 63.9 ± 18.2 | 58.4 ± 18.5 | 58.5 ± 20.8 | < 0.0001 |
| **NT-pro BNP, pg/ml** | 440 (908, 119) | 831 (416, 1592) | 979 (550, 2022) | 0.0040 |
| **BNP, pg/ml** | 95 (39, 109) | 195 (108, 350) | 187 (92, 285) | 0.010 |
| **DR-FLASH score** | 3.4 ± 1.2 | 4.2 ± 1.2 | 4.3 ± 1.1 | < 0.0001 |
| **SPEED score** | 2.0 ± 1.1 | 3.0 ± 1.0 | 3.2 ± 1.0 | < 0.0001 |
| **Echocardiography findings** |  |  |  |  |
| *Left atrial diameter, mm* | 40.1 ± 6.8 | 41.8 ± 6.7 | 42.7 ± 6.4 | < 0.0001 |
| *Left ventricular ejection fraction, %* | 61.4 ± 12.0 | 60.5 ± 12.5 | 60.6 ± 14.0 | 0.574 |

AF, atrial fibrillation; GFR, glomerular filtration rate; NT-pro BNP, N-terminal prohormone of brain natriuretic peptide; BNP, brain natriuretic peptide.

Procedural characteristics are shown in Table 2. Fluoroscopic time was longer in patients with LVAs. There were differences in the distribution of 3-dimensional mapping systems and ablation catheters used for pulmonary vein isolation. As LVA size became more extensive, the frequency of non-pulmonary-vein trigger, roof line, bottom line, and LVA ablation became higher.

**Table 2.**
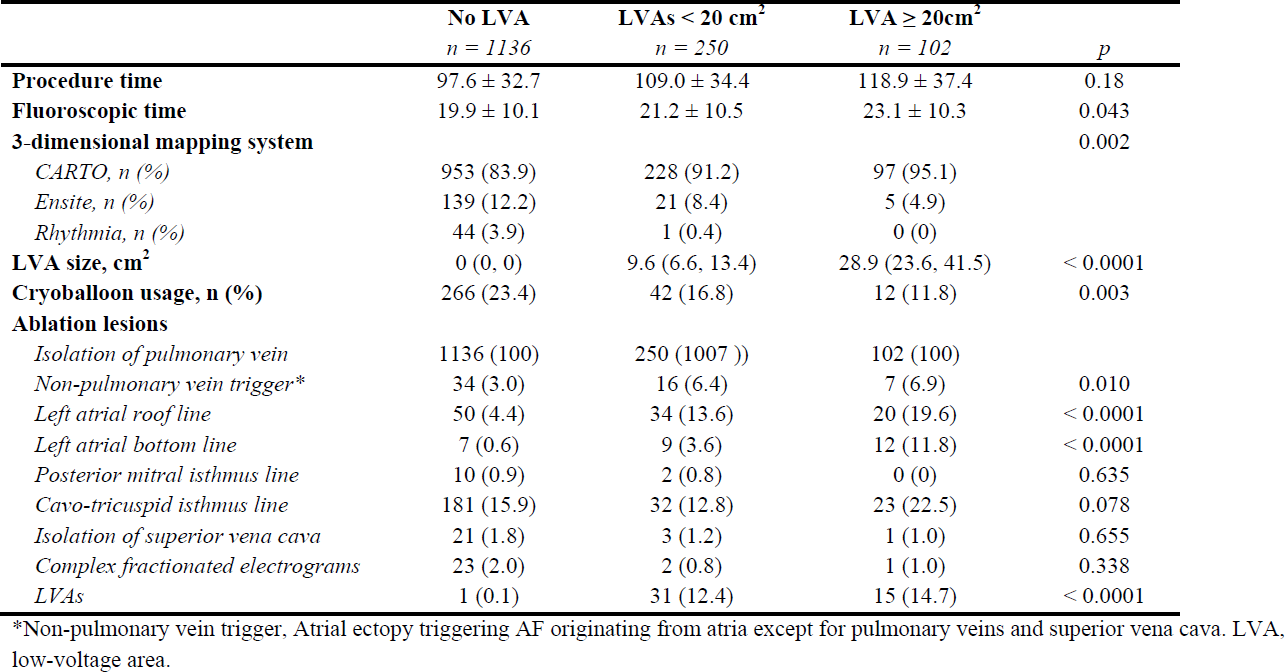
Procedure characteristics.

|  | No LVA<br>*n* = 1136 | LVAs < 20 cm<sup>2</sup><br>*n* = 250 | LVA ≥ 20cm<sup>2</sup><br>*n* = 102 | *p* |
| --- | --- | --- | --- | --- |
| **Procedure time** | 97.6 ± 32.7 | 109.0 ± 34.4 | 118.9 ± 37.4 | 0.18 |
| **Fluoroscopic time** | 19.9 ± 10.1 | 21.2 ± 10.5 | 23.1 ± 10.3 | 0.043 |
| **3-dimensional mapping system** |  |  |  | 0.002 |
| *CARTO, n (%)* | 953 (83.9) | 228 (91.2) | 97 (95.1) |  |
| *Ensite, n (%)* | 139 (12.2) | 21 (8.4) | 5 (4.9) |  |
| *Rhythmia, n (%)* | 44 (3.9) | 1 (0.4) | 0 (0) |  |
| **LVA size, cm<sup>2</sup>** | 0 (0, 0) | 9.6 (6.6, 13.4) | 28.9 (23.6, 41.5) | < 0.0001 |
| **Cryoballoon usage, n (%)** | 266 (23.4) | 42 (16.8) | 12 (11.8) | 0.003 |
| **Ablation lesions** |  |  |  |  |
| *Isolation of pulmonary vein* | 1136 (100) | 250 (100%) | 102 (100) |  |
| *Non-pulmonary vein trigger\** | 34 (3.0) | 16 (6.4) | 7 (6.9) | 0.010 |
| *Left atrial roof line* | 50 (4.4) | 34 (13.6) | 20 (19.6) | < 0.0001 |
| *Left atrial bottom line* | 7 (0.6) | 9 (3.6) | 12 (11.8) | < 0.0001 |
| *Posterior mitral isthmus line* | 10 (0.9) | 2 (0.8) | 0 (0) | 0.635 |
| *Cavo-tricuspid isthmus line* | 181 (15.9) | 32 (12.8) | 23 (22.5) | 0.078 |
| *Isolation of superior vena cava* | 21 (1.8) | 3 (1.2) | 1 (1.0) | 0.655 |
| *Complex fractionated electrograms* | 23 (2.0) | 2 (0.8) | 1 (1.0) | 0.338 |
| *LVAs* | 1 (0.1) | 31 (12.4) | 15 (14.7) | < 0.0001 |

\*Non-pulmonary vein trigger, Atrial ectopy triggering AF originating from atria except for pulmonary veins and superior vena cava. LVA, low-voltage area.

### Prognostic impact of LVAs

Composite endpoints developed in 105 (7.1%) of 1,488 patients, and AF recurrence occurred in 410 (27.6%). Figure 1A demonstrates that composite endpoints developed more frequently in patients with a larger LVA size in the order LVA size of ≥ 20 cm^2^, < 20 cm^2^ and no LVAs. AF recurrence was more often observed in patients with a larger LVA size in the order LVA size of ≥ 20 cm^2^, < 20 cm^2^ and no LVAs (Figure 2). The finding of higher frequencies of composite endpoints in patients with LVAs than in those without LVAs was consistent, irrespective of the presence or absence of AF recurrence (Figure 1B and 1C). However, the tendency toward more frequent composite endpoints in patients with extensive LVAs (≥ 20 cm^2^) than in those with small LVAs (< 20cm^2^) observed in the sub-analysis of patients with AF recurrence was not seen in that of patients without AF recurrence. Annual incidence rates of each endpoint are shown in Figure 3. Each endpoint demonstrated higher frequency in patients with LVAs than in those without LVAs.

**Figure 1.**
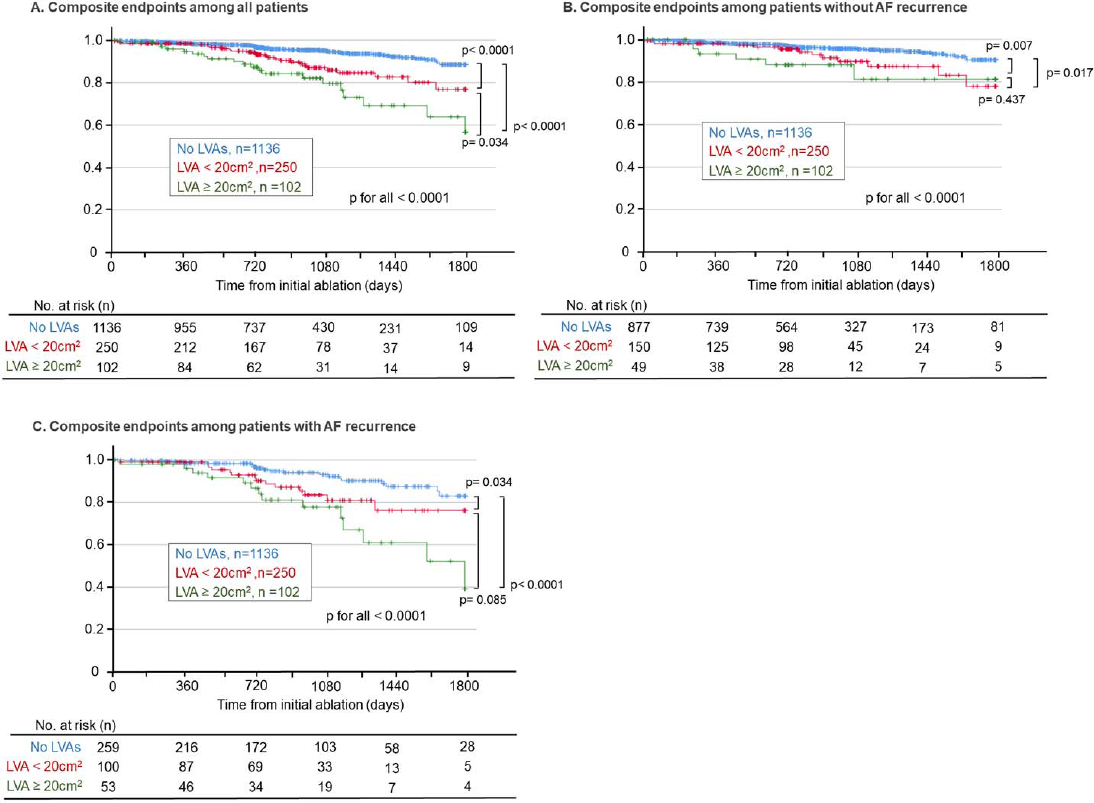
Kaplan-Meier curves of composite-endpoint*-free rates. Kaplan-Meier curves of the composite-endpoint-free rate after initial ablation. Comparison was performed between patients with no LVA, those with small LVAs (< 20cm^2^), and those with extensive LVAs (≥ 20cm^2^). Composite endpoints developed more frequently in the order of patients with extensive LVAs, small LVAs, and no LVA among all patients (A). Patients with LVAs experienced composite endpoints more frequently than those without LVAs, irrespective of with or without AF recurrence (B and C). *Composite endpoints included death, heart failure, and stroke. LVA indicates low-voltage area.

**Figure 2.**
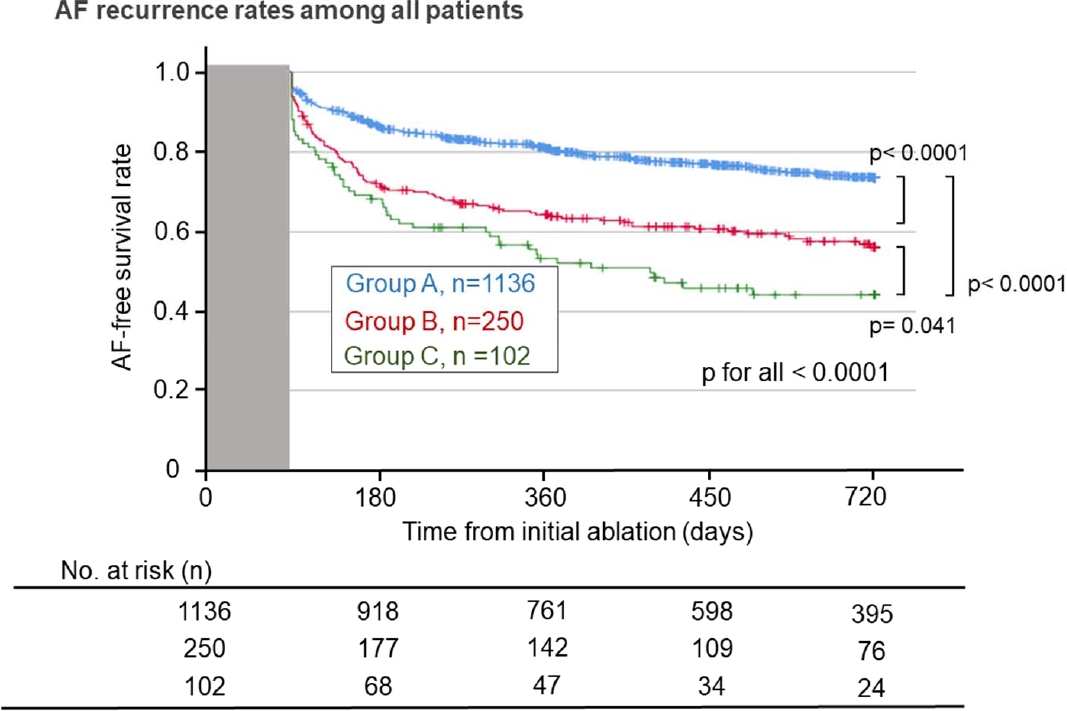
Kaplan-Meier curves of AF recurrence-free rates. Kaplan-Meier curves of AF recurrence-free rate after initial ablation. Comparison was performed between patients with no LVA, those with small LVAs (< 20cm^2^), and those with extensive LVAs (≥ 20cm^2^). AF recurrence developed more frequently in the order of patients with extensive LVAs, small LVAs, and no LVA. LVA indicates low-voltage area.

**Figure 3.**
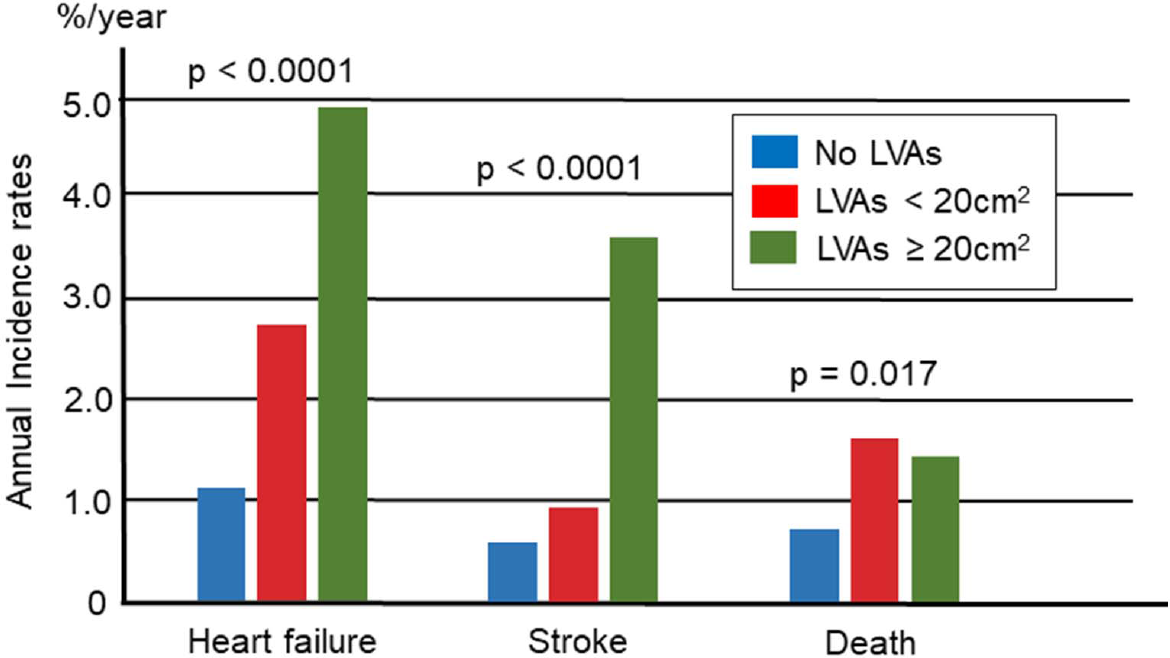
Annual incidence of heart failure, stroke, and death. Annual incidence of heart failure, stroke, and death stratified by LVA size in all patients (A), patients without AF recurrence (B), and patients with AF recurrence (C) are presented. Patients with LVAs showed higher incidence of heart failure, stroke, and death in all patients (A). LVA indicates low-voltage area.

### Death and LVAs

In total, 34 (2.3%) patents died, with 9 (0.6%) due to cardiovascular causes (Table 3). These cardiovascular deaths consisted of 7 cases of heart failure, 1 of thromboembolism of the superior mesenteric artery, and 1 of bacterial endocarditis. Cardiovascular deaths were more frequent in patients with LVAs than in those without. In contrast, no difference was observed in the frequencies of non-cardiovascular deaths between those with and without LVAs.

**Table 3.**
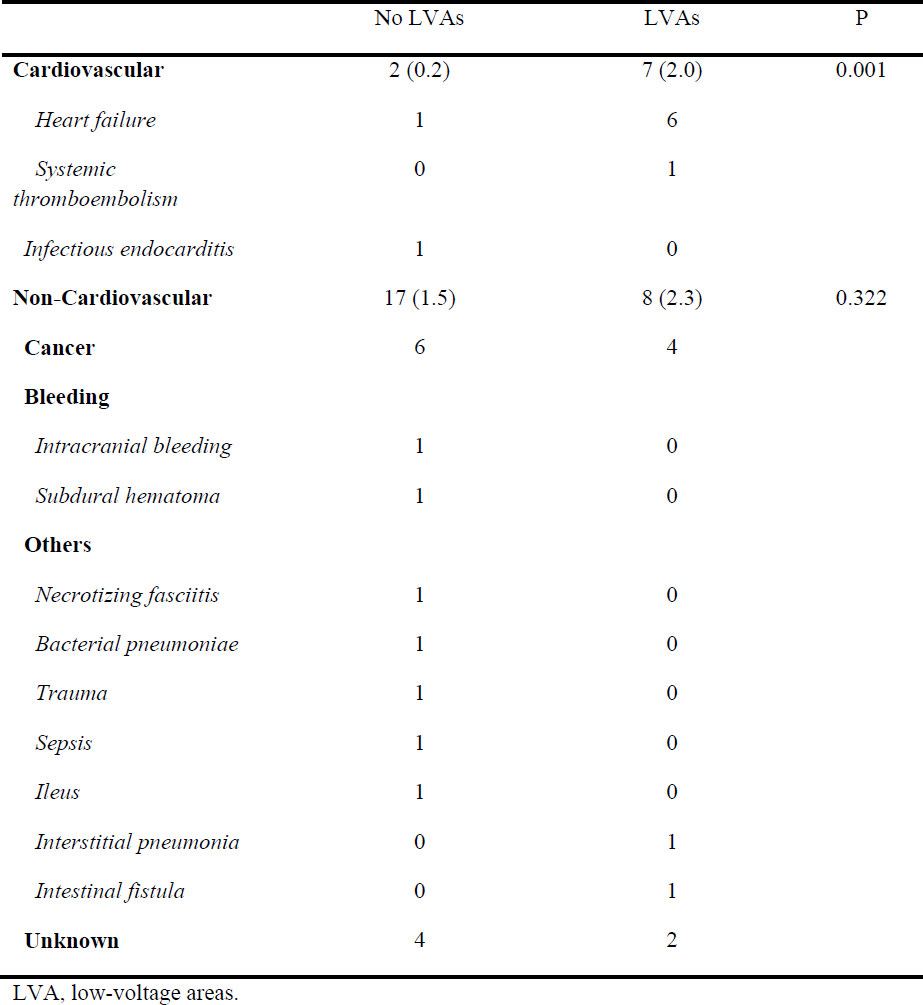
Causes of death.

### Multivariable analyses for prediction of composite endpoints

Univariable and multivariable analyses revealed that LVA presence was independently associated with the development of composite endpoints (Table 4). Other independent factors related to composite endpoints were concomitant diabetes mellitus, history of heart failure, low estimated glomerular filtration rate, large left atrial diameter, and AF recurrence.

**Table 4.** Factors associated with composite endpoints.

|  | **Composite endpoints** |  | **Univariate** |  |  | **Multivariate** |  |  |
| --- | --- | --- | --- | --- | --- | --- | --- | --- |
|  | *With*<br>*n = 105* | *Without*<br>*n = 1383* | *HR* | *95% CI* | *p* | *HR* | *95% CI* | *p* |
| **Age, years** | 70.5 ± 9.4 | 68.0 ± 10.2 | 1.03 | 1.01-1.05 | 0.007 | 1.01 | 0.98-1.03 | 0.67 |
| **Female, n (%)** | 40 (38.1) | 461 (33.3) | 1.21 | 0.82-1.80 | 0.338 |  |  |  |
| **Body mass index** | 23.5 ± 4.4 | 24.1 ± 4.0 | 0.96 | 0.92-1.02 | 0.162 |  |  |  |
| **Persistent AF, n (%)** | 66 (62.9) | 839 (60.7) | 1.30 | 0.87-1.93 | 0.195 |  |  |  |
| **Hypertension, n (%)** | 56 (53.3) | 778 (56.3) | 0.89 | 0.61-1.31 | 0.561 |  |  |  |
| **Diabetes mellitus, n (%)** | 31 (29.5) | 238 (17.2) | 2.14 | 1.41-3.25 | <0.0001 | 1.82 | 1.19-2.79 | 0.006 |
| **Heart failure, n (%)** | 57 (54.3) | 265 (19.2) | 4.78 | 3.25-7.02 | <0.0001 | 3.37 | 2.26-5.01 | <0.0001 |
| **Stroke/thromboembolism, n (%)** | 9 (8.6) | 108 (7.8) | 1.07 | 0.54-2.11 | 0.852 |  |  |  |
| **Vascular disease, n (%)** | 17 (16.2) | 117 (8.5) | 2.11 | 1.26-3.55 | 0.005 | 1.13 | 0.65-1.97 | 0.66 |
| **eGFR, mL/min/1.73m<sup>2</sup>** | 48.5 ± 22.6 | 63.7 ± 17.8 | 0.97 | 0.96-0.97 | <0.0001 | 0.97 | 0.96-0.98 | <0.0001 |
| **AF recurrence** | 47 (44.8) | 365 (26.4) | 2.05 | 1.40-3.02 | <0.0001 | 1.85 | 1.24-2.77 | 0.003 |
| **LVA presence, n (%)** | 47 (44.8) | 305 (22.1) | 2.86 | 1.94-4.20 | <0.0001 | 1.73 | 1.13-2.64 | 0.011 |

Factors with $p < 0.05$ in the univariate analysis were incorporated in the multivariate analysis. AF, atrial fibrillation; eGFR, estimated glomerular filter rate; LVA, low-voltage area.

## Discussion

This observational study compared the composite endpoints of death, heart failure, and stroke in patients who underwent initial AF ablation with stratification by left atrial LVA size. The main findings were as follows. First, composite endpoints were more frequent in the order of patients with extensive LVAs (≥ 20 cm^2^), those with small LVAs (< 20 cm^2^) and those without LVAs. Second, the annual incidence of each endpoint (death, heart failure, and stroke) was higher in patients with LVAs than in those without LVAs. The presence of LVAs was independently associated with higher incidence of composite endpoints; notably, this association was irrespective of AF recurrence. To our knowledge, this is the first study to report a negative prognostic impact of atrial cardiomyopathy as assessed by size of left atrial LVAs.

### Left atrial LVAs and poor prognosis

For many decades, AF was thought to be associated with death, stroke, and heart failure. Recently published reports proposed a new term, atrial cardiomyopathy, to indicate the advanced atrial remodeling underlying AF development.^2^ Atrial cardiomyopathy is an essential pathophysiological characteristic of AF and is reportedly associated with stroke irrespective of AF, through loss of atrial function and injury to the atrial endocardium.^15^

Recent clinical studies suggest that atrial cardiomyopathy also contributes to the development of heart failure in AF patients. In addition to conventional contributors to heart failure concomitant with AF, such as loss of atrial contraction and irregular and fast ventricular beats, cardiac function is also damaged by factors related to atrial cardiomyopathy, including stiff left atria,^4^ functional mitral regurgitation,^16^ and loss of secretion of atrial natriuretic peptides.^17^

Conversely, advanced atrial cardiomyopathy might also be explained as a result of heart or systemic conditions with poor prognosis, and LVAs might act as markers which predict poor prognosis. Under this scenario, for example, left ventricular dysfunction might cause left atrial remodeling through the elevation of left atrial pressure.^18, 19^ In addition, systemic conditions such as aging might predispose to atrial myocardial degeneration.^20,21^

In the sub-analyses of patients stratified by the presence or absence of AF recurrence, the prognostic impact of LVA and its extension was more obvious among patients with AF recurrence than in those without. As shown in Table 3, both LVA and AF recurrence were associated with composite endpoints, independently of each other. This finding suggests the existence of multiple different pathophysiological pathways leading to the composite endpoints, and that LVA and AF recurrence may have a synergistic impact on development of the composite endpoints.

### Prediction of poor prognosis using other clinical parameters

Notably, other parameters representing advanced atrial remodeling, large left atrial diameter and AF recurrence were also independently associated with poor prognosis. These results suggest that atrial remodeling is a multiform pathological process. Accordingly, the association between atrial remodeling and poor prognosis is likely subject to significant inter-individual differences.

In addition to left atrial parameters indicating advanced left atrial remodeling, we also found that factors outside the left atrium such as concomitant diabetes mellitus, history of heart failure, and reduced kidney function were associated with poor prognosis. Previous studies have reported that patients with these factors have high risk of stroke^22–24^ and heart failure.^25,26^ Mortality might also be increased due to increased risk of cancer in patients with diabetes mellitus, heart failure, and chronic kidney disease,^27–29^ in addition to cardiovascular disease-related death.

### Clinical implications

In this study, advanced atrial cardiomyopathy, as represented by the presence of LVA and its extension, was shown to be an independent predictor of the composite endpoints of death, stroke, and heart failure. These results suggest that management of AF patients should take account of atrial cardiomyopathy. Future studies are expected to investigate whether medications which act to suppress ventricular remodeling, such as inhibitors of the renin-angiotensin-system, sodium-glucose cotransporter-2 inhibitors, and mineralocorticoid antagonists, are also effective in suppressing atrial remodeling.^30^ In addition, continuation of oral anticoagulants aimed at preventing ischemic stroke might be preferable even when sinus rhythm is maintained after ablation.

### Limitations

Several limitations of this study warrant mention. First, the study was conducted under a retrospective design at a single institution, and might accordingly have been subject to a variety of biases. Second, small LVAs (< 5cm^2^) were not considered diagnostic of the presence of LVA in order to avoid misjudgment. Third, voltage maps and LVAs were possibly influenced by the different mapping catheters and different mapping systems used. These factors can influence the results of a voltage map. Fourth, selection of ablation procedures was at the discretion of the operator, albeit based on international guidelines and electrophysiological findings, and were accordingly not necessarily consistent. This might have biased the AF recurrence rate. Fifth, AF recurrence after discharge was quantified on the basis of the patient’s symptoms, giving rise to the possibility that asymptomatic episodes of AF might have been missed. Sixth, AF recurrence was followed for 2 years after the initial ablation, and cardiac rhythm after that period or after repeat ablation were not considered, possibly influencing the results. Seventh, in some cases, information on 5-year composite endpoints was collected by face-to-face or telephone interview without confirmation using objective medical records.

### Conclusion

LVA presence and its extent were associated with poor long-term composite endpoints including death, heart failure, and stroke, irrespective of AF recurrence and other confounders. The results suggest that underlying atrial cardiomyopathy indicates poor prognosis after AF ablation. Atrial cardiomyopathy should be given greater attention in the management of patients with AF, particularly when atrial remodeling is advanced.

## Source of funding

None

## Disclosures

Masuda M has received honoraria from Medtronic, Johnson & Johnson, Boston Scientific, Abbott, and Nihon Kohden outside the submitted work. Ishihara T has received honoraria from Nipro and Kaneka Medics outside the submitted work.

## Data Availability

Data will be available in response to request by email.

## Notes

### Competing Interest Statement

The authors have declared no competing interest.

### Funding Statement

No external funding

### Author Declarations

IRB in Kansai Rosai Hospital

